## Supplementary Figures and Tables for "SARS-CoV-2 and other respiratory pathogens are detected in continuous air samples from congregate settings"

### Supplementary Information

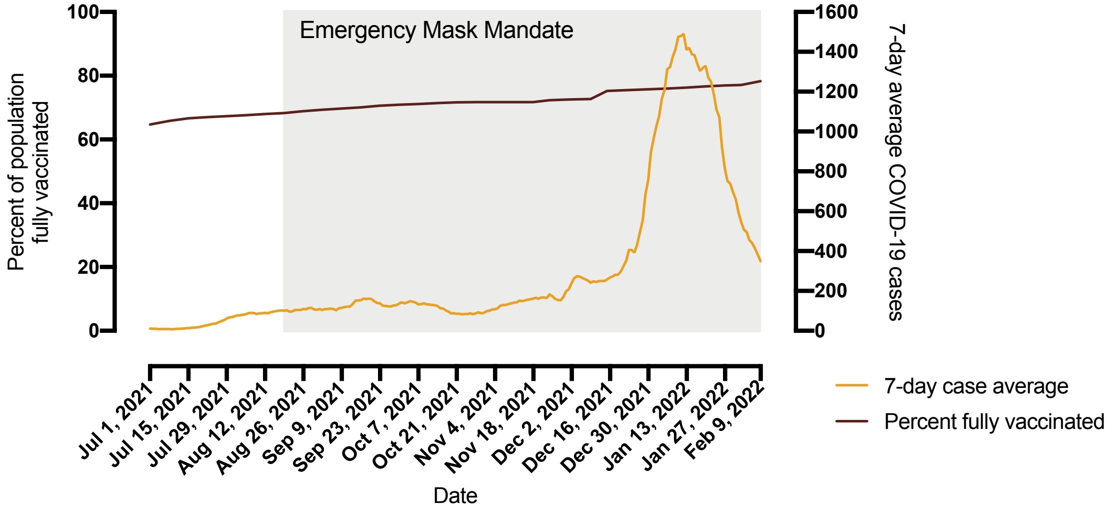


**Supplementary Figure 01. COVID-19 seven-day average cases and vaccination rates in Dane County, WI.** The percent of fully vaccinated individuals (red) is plotted on the left y-axis and seven-day average COVID-19 cases (orange) in Dane County, WI is plotted on the right y-axis with respect to time on the x-axis. Data were downloaded from Public Health Madison and Dane County (PHMDC) coronavirus dashboard (https://publichealthmdc.com/coronavirus/dashboard; accessed on March 3, 2022). Seven-day average COVID-19 cases were calculated from the sum of new cases per day for the most recent seven-day period divided by seven and rounded to the nearest whole number. The percent of the population fully vaccinated was calculated from the total number of residents who received all recommended doses in the primary series of the respective vaccines, this excludes additional recommended booster doses. The gray box shows the dates that a face-covering emergency order was implemented in Dane County, WI by PHMDC.

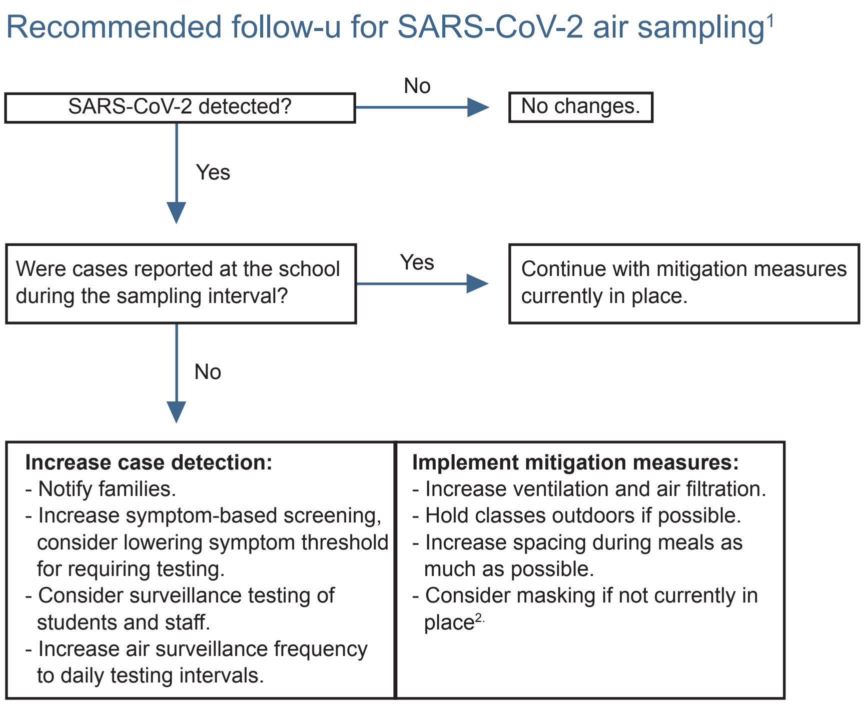


**Supplementary Figure 02. SARS-CoV-2 air sampling response flowchart.** Air sampling response decision flowchart was developed in collaboration with Public Health Madison and Dane County (PHMDC) to describe how schools may use air surveillance to implement risk mitigation strategies. [1] These recommendations supplement but would not supersede universal recommendations for all schools from PHMDC. [2] Universal masking recommended for all teachers, students, and staff in K-12 settings and may be required by public health orders.

#### Supplementary Table 1. TrueMark Respiratory Panel 2.0 TaqMan Array Card Crt cut-off values.

| Target | Pathogen name (what is listed in Fig 4) | Minimum number of copies per reaction | Mean Crt | Comments |
|---|---|---|---|---|
| AdV_2of2-Vi99990002_po | Adenovirus | 1.25 | 34.5 | Adenovirus was called positive if AdV_1of2 or AdV_2of2 were positive. |
| B.pertussis-Ba06439623_s1 | Bordetella spp. | 1.25 | 34.63 | Bordetella spp. was called positive if one of these targets were positive. |
| Bordetella-Ba06439624_s1 | Bordetella spp. | 1.25 | 31.93 | |
| C.pneumoniae-Ba06439616_s1 | Chlamydophila pneumoniae | 50 | 33.41 | |
| CoV_229E-Vi06439671_s1 | Human coronavirus 299E | 50 | 35.13 | |
| CoV_NL63-Vi06439673_s1 | Human coronavirus NL63 | 250 | 33.12 | |
| CoV_OC43-Vi06439646_s1 | Human coronavirus OC43 | 50 | 34.99 | |
| EV_D68-Vi06439669_s1 | Human enterovirus | 1.25 | 33.11 | Human enterovirus was called positive if EV_D6 or EV_pan were positive. |
| Flu_A_H1-Vi99990009_po | Influenza A virus | 12.5 | 31.59 | Influenza A virus was called positive if one of these targets were positive. |
| Flu_A_H3-Vi99990010_po | Influenza A virus | 1.25 | 35.71 | |
| Flu_A_pan-Vi99990011_po | Influenza A virus | 1.25 | 33.89 | |
| H.influenzae-Ba06439625_s1 | Haemophilus influenzae | 250 | 35.03 | |
| HBoV-Vi99990003_po | Human bocavirus | 12.5 | 32.58 | |
| HHV3-Vi06439647_s1 | Varicella zoster virus | 12.5 | 35.7 | |
| HHV4-Vi06439675_s1 | Epstein-Barr virus | 1.25 | 34.1 | |
| HHV5-Vi06439643_s1 | Cytomegalovirus | 1.25 | 33.77 | |
| hMPV-Vi99990004_po | Metapneumovirus | 12.5 | 34.73 | |
| hPIV1-Vi06439642_s1 | Parainfluenza virus | 50 | 33.94 | Parainfluenza virus was called positive if one of these targets were positive. |
| hPIV2-Vi06439672_s1 | Parainfluenza virus | 12.5 | 34.08 | |
| hPIV3-Vi06439670_s1 | Parainfluenza virus | 12.5 | 33.75 | |
| hPIV4-Vi99990005_po | Parainfluenza virus | 250 | 32.96 | |
| K.pneumoniae-Ba04932083_s1 | Klebsiella pneumoniae | 1.25 | 34.16 | |
| L.pneumophila-Ba06439617_s1 | Legionella pneumophila | 250 | 33.63 | |
| M.catarrhalis-Ba06439622_s1 | Moraxella catarrhalis | 12.5 | 35.4 | |
| M.pneumoniae-Ba06439620_s1 | Myoplasma pneumoniae | 12.5 | 27.97 | |
| Measles -Vi99990013_po | Measles virus | 1.25 | 35.3 | |
| MERS_CoV-Vi06439644_s1 | MERS | 50 | 34.96 | |
| Mumps-Vi06439657_s1 | Mumps virus | 12.5 | 30.01 | |
| RSVA-Vi99990014_po | RSVA | 12.5 | 36.58 | |
| RSVB -Vi99990015_po | RSVB | 12.5 | 33.61 | |
| RV_1of2-Vi99990016_po | Human rhinovirus | 50 | 35.35 | Human rhinovirus was called positive if RV_1of2 or RV_2of2 were positive. |
| S.pneumoniae-Ba06439619_s1 | Streptococcus pneumoae | 250 | 33.67 | |
| HPeV-Vi99990006_po | Human parechovirus | 1.25 | 33.7 | |
| **Not detectable at less than 250 copies per reaction but included in Fig. 4 if Crt<30** | | | | |
| CoV_HKU1-Vi06439674_s1 | Human coronavirus HKU1 | NA | 30 | |
| EV_pan-Vi06439631_s1 | Human enterovirus | NA | 30 | Human enterovirus was called positive if EV_D6 or EV_pan were positive. |
| Flu_B_pan-Vi99990012_po | Influenza B virus | NA | 30 | |
| RV_2of2-Vi99990017_po | Human rhinovirus | NA | 30 | Human rhinovirus was called positive if RV_1of2 or RV_2of2 were positive. |
| **Detected in isolated pooled air cartridges but included in Fig. 4 if Crt<30** | | | | |
| AdV_1of2-Vi99990001_po | Adenovirus | NA | 30 | Adenovirus was called positive if AdV_1of2 or AdV_2of2 were positive |
| S.aureus-Ba04646259_s1 | Staphylococcus aureus | NA | 30 | |

**Supplementary Table 2. Respiratory pathogens detected in air samples collected from congregate settings.**

| Category | Pathogen | Count |
|---|---|---|
| Acute Viruses | Adenovirus | 6 |
| | Human coronavirus OC43 | 7 |
| | Influenza A virus | 30 |
| | Influenza A virus H3 subtype | 19 |
| | Human bocavirus | 7 |
| | Human parainfluenza virus (hPIV3) | 1 |
| | Respiratory syncytial virus A (RSVA) | 2 |
| | Respiratory syncytial virus B (RSVB) | 1 |
| Persistent Viruses | Varicella zoster virus (HHV3) | 1 |
| | Epstein-Barr Virus (HHV4) | 40 |
| | Cytomegalovirus (HHV5) | 36 |
| Commensal Bacteria | Haemophilus influenzae | 1 |
| | Klebsiella pneumoniae | 96 |
| | Moraxella catarrhalis | 26 |
| | Staphylococcus aureus | 93 |
| | Streptococcus pneumoniae | 5 |
| Atypical bacteria | Bordetella spp. | 2 |
